# Spectral performance evaluation of a second-generation spectral detector CT

**DOI:** 10.1101/2022.06.03.22275935

**Authors:** Leening P. Liu, Nadav Shapira, Sandra S. Halliburton, Sebastian Meyer, Amy Perkins, Harold I. Litt, Hans Ulrich Kauczor, Tim Leiner, Wolfram Stiller, Peter B. Noël

**Affiliations:** Department of Radiology, Perelman School of Medicine, University of Pennsylvania, Philadelphia, PA, USA; Department of Bioengineering, University of Pennsylvania, Philadelphia, PA, USA; Philips Healthcare, Orange Village, OH, USA; Diagnostic and Interventional Radiology (DIR), Heidelberg University Hospital, Heidelberg, Germany; Department of Radiology, Mayo Clinic, Rochester, MN, USA

**Keywords:** x-ray computed tomography, diagnostic imaging, multidetector computed tomography

## Abstract

**Objective:** To characterize a second-generation wide-detector dual-layer spectral computed tomography system for material quantification accuracy, acquisition parameter and patient size dependencies, and tissue characterization capabilities.

**Methods:** A phantom with multiple tissue-mimicking and material inserts was scanned with a dual-layer spectral detector CT using different tube voltages, collimation widths, radiation dose levels, and size configurations. Accuracy of iodine density maps and virtual monoenergetic images (MonoE) were investigated. Additionally, differences between conventional and MonoE 70 keV images were calculated to evaluate acquisition parameter and patient size dependencies. To demonstrate material quantification and differentiation, liver-mimicking inserts with adipose and iron were analyzed with a two-base decomposition utilizing MonoE 50 and 150 keV, and root mean square error (RMSE) for adipose and iron content was reported.

**Results:** Spectral accuracy was high for the measured inserts across a wide range of MonoE levels. MonoE 70 keV demonstrated reduced dependence compared to conventional image for phantom size (3 vs. 29 HU) and acquisition parameters, particularly tube voltage (5 vs. 43 HU) and noise-dose (9 vs. 11 HU). Iodine density quantification was successful with errors ranging from 0.25 to 0.5 mg/mL. Similarly, inserts with different amounts of adipose and iron were easily differentiated, and the small deviation in values within inserts corresponded to a RMSE of 1.78 ± 0.37% and 0.87 ± 0.37 mg/mL for adipose and iron content, respectively.

**Conclusion:** The second-generation dual-layer CT enables acquisition of quantitatively accurate spectral data without compromises from differences in patient size and acquisition parameters.

**Key Points:**

- With second-generation wide-detector dual-layer computed tomography, spectral quantification is further improved.
- Spectral performance is independent of acquisition parameters such as tube voltage (100 kVp versus 120 kVp) and z-coverage (10 versus 80 mm).
- Spectral performance is not significantly impacted by patient habitus.

## Introduction

Computed Tomography (CT) is often one of the first imaging studies that a patient undergoes when entering the healthcare system and is part of the surveillance workflow for various pathologies and disease states. However, conventional CT still poses longstanding challenges for the clinical day-to-day routine. Traditionally, CT utilizes a polyenergetic x-ray beam and energy-integrating detectors to produce cross-sectional image data based on attenuation measured in Hounsfield Units (HU). In certain situations, i.e. kidney stones [1, 2] and liver lesions [3], different elements or tissues can be challenging to distinguish based only on HU values. Conventional CT is also not able to measure physical quantities, e.g. iodine density in units of mg/mL. Conventional CT image quality can additionally be negatively affected by patient habitus.

In contrast, spectral CT, as dual-energy CT (DECT) or multi-energy CT, enables differentiation and quantitative characterization of elements or tissues [4, 5]. It relies on the fact that different elements exhibit different levels of attenuation when exposed to x-ray photons of different energies from a polyenergetic x-ray source. The attenuation is attributed to two main effects: the photoelectric effect and Compton scattering. The photoelectric effect depends on the atomic number and dominates at energies up to 100 keV, while Compton scattering is largely dependent on physical density and dominates at higher energies [6]. There are several different technical approaches to obtaining spectral CT information, including dual-source [7], split-filter [8], kVp-switching [9, 10], dual-layer [11, 12], and photon-counting CT [13, 14]. The various technological solutions make it possible to obtain spectral data of varying quality [15–17], and generally there is a trend towards detector-based solutions.

Dual-layer spectral detector CT allows routine acquisition of clinical DECT data without special acquisition protocols [18]. It features a dual-layer detector that separates low energy data from high energy data. The top layer detects low-energy photons, while the bottom layer detects high-energy photons. Projection data obtained simultaneously from both detector layers can then be utilized to generate spectral images in addition to conventional CT image data. This technology allows spectral data to be acquired on all studies for all patients without selecting any special “spectral protocols” beforehand.

For almost all organ systems, availability of spectral data frequently adds valuable information for CT interpretation [19–24]. The use of virtual monoenergetic images (MonoE) at low energies can improve contrast signal in vascular studies, including those with suboptimal enhancement [25, 26]. MonoEs at high energies, on the other hand, can reduce artifacts, particularly those from implants and stents, to reveal information previously hidden by artifacts [27–29]. With accurate iodine measurements from iodine density images, perfusion deficits can be identified, thereby improving diagnostics for pulmonary embolism [30, 31], myocardial infarction [32–34], and oncological lesions [35–38], for instance. In these applications and others, an increased z-coverage with wider detectors has additional advantages including a reduction in acquisition time and larger anatomical coverage of the tissue(s) or organ(s) of interest per gantry rotation.

This study reports on the technical performance of a dual-layer CT (Spectral CT 7500, Philips Healthcare) with a z-coverage of 8 cm and an 80 cm bore. The system is equipped with a second-generation dual-layer detection system, which consists of a higher-efficiency detector array coupled with a two-dimensional anti-scatter grid. In comparison, the previous generation detector array was a stick design, which read out the scintillator on one side and limited by a one-dimensional anti-scatter grid. Our evaluation highlights the spectral performance of the second-generation system in regards to material quantification accuracy, tissue characterization capabilities, and acquisition parameter and patient size dependencies.

## Methods

### Phantom

A phantom with interchangeable material inserts (Multi-energy CT Phantom, Gammex, Sun Nuclear) was utilized (Figure 1A). It consisted of an extension ring (30 × 40 cm, ‘large’) and an inner cylindrical portion with a diameter of 20 cm (‘small’). The interchangeable cylindrical inserts included tissue-mimicking inserts as well as inserts containing a known concentration of an element, e.g. iodine. We calculated the expected attenuation at selected energies (keVs) and expected iodine densities as ground truth for comparison with measured values using the material composition and insert density information supplied by the phantom manufacturer [39]. The manufacturer provided values for adipose content and iron densities were utilized as ground truth for adipose and iron quantification. For examining spectral accuracy and acquisition parameter and patient size dependence, a specific insert configuration (config 1, Figure 1B) was utilized. It included inserts for 2 mm iodine core, 5 mm iodine core, adipose, blood 40, blood 70, blood 110, blood + 2 mg/mL iodine, blood + 4 mg/mL iodine, brain, calcium 50 mg/mL, calcium 100 mg/mL, calcium 300 mg/mL, iodine 2 mg/mL, iodine 5 mg/mL, iodine 10 mg/mL, and iodine 15 mg/mL. For assessing material characterization, a different insert configuration (config 2, Figure 1C) was scanned and featured inserts for adipose, blood 40, blood 70, blood 110, brain grey matter, brain white matter, iodine 5 mg/mL, liver + 10% adipose, liver + 20% adipose, liver + 35% adipose, liver + 50% adipose, liver + 5 mg/mL iron, liver + 8 mg/mL iron, liver + 11 mg/mL iron, and solid water.

**Figure 1.**
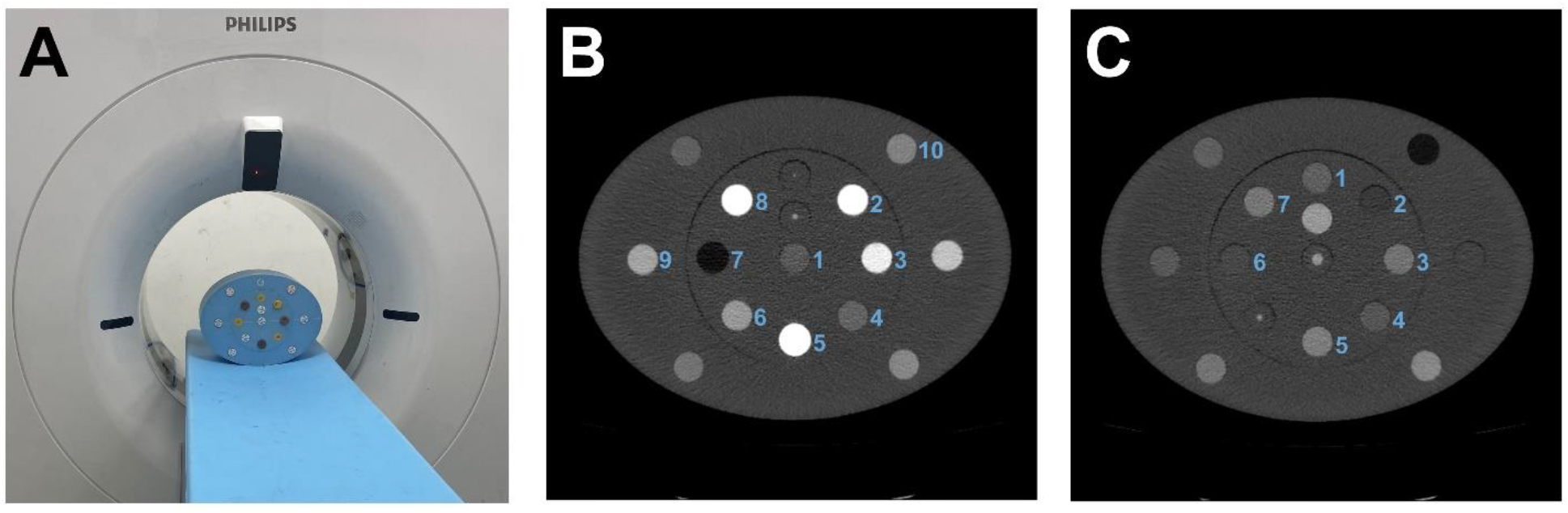
Insert configurations for the evaluation of spectral accuracy, quantitative stability, and material characterization. (A) The phantom was scanned on a second-generation dual-layer spectral detector CT. (B) For the assessment of accuracy and stability, relevant inserts included: 1. brain, 2. calcium 100 mg/mL, 3. iodine 10 mg/mL, 4. iodine 2 mg/mL, 5. calcium 300 mg/mL, 6. iodine 5 mg/mL, 7. adipose, 8. iodine 15 mg/mL, 9. blood + 4 mg/mL iodine, and 10. blood + 2 mg/mL iodine. (C) For demonstrating material separation with spectral results, inserts included: 1. liver + 10% adipose, 2. liver + 50% adipose, 3. liver + 5 mg/mL iron, 4. liver + 20% adipose, 5. liver + 11 mg/mL iron, 6. liver + 35% adipose, and 7. liver + 8 mg/mL iron.

### Image acquisition

The phantom was scanned on a second-generation dual-layer spectral detector CT. Scans were performed at 100 and 120 kVp for both phantom sizes (large, small). In addition, collimation width (16×0.625, 128×0.625 mm) and noise index/dose right index (DRI 10, 16, 23) were varied. The resulting volumetric CT dose index (CTDI_vol_) for each combination of parameters is listed in Table 1. Scans were repeated three times for each insert configuration, phantom size, tube voltage, collimation width, and DRI level combination to capture the statistics of reconstructed results. Other relevant parameters are presented in Table 2. Spectral results were reconstructed for analysis, specifically iodine density maps, and MonoEs at 40, 50, 60, 70, 100, 150, and 200 keV. Conventional CT images were also reconstructed to serve as a comparison to the MonoE 70 keV. Unlike other dual-energy technologies, the dual-layer technology is able to generate true conventional images by integrating data from the lower and upper layers.

**Table 1.** Corresponding radiation dose in terms of volumetric CT dose index (CTDI_vol_) for combinations of tube voltage, phantom size, and collimation.

| Tube Voltage [kVp] | Size | Collimation [mm] | DRI 10 |  | DRI 16 |  | DRI 23 |  |
| --- | --- | --- | --- | --- | --- | --- | --- | --- |
|  |  |  | Exposure [mAs] | CTDI<sub>vol</sub> [mGy] | Exposure [mAs] | CTDI<sub>vol</sub> [mGy] | Exposure [mAs] | CTDI<sub>vol</sub> [mGy] |
| 100 | large | 16x0.625 | 106 | 7.8 | 167 | 12.2 | 270 | 19.8 |
| 100 | large | 128x0.625 | 106 | 5.1 | 167 | 8 | 270 | 13 |
| 100 | small | 16x0.625 | 25 | 1.8 | 39 | 2.9 | 107 | 7.8 |
| 100 | small | 128x0.625 | 25 | 1.2 | 39 | 1.9 | 107 | 5.1 |
| 120 | large | 16x0.625 | 64 | 7.6 | 101 | 11.9 | 270 | 31.9 |
| 120 | large | 128x0.625 | 64 | 5 | 101 | 7.8 | 270 | 20.9 |
| 120 | small | 16x0.625 | 16 | 1.9 | 25 | 3 | 69 | 8.1 |
| 120 | small | 128x0.625 | 16 | 1.2 | 25 | 1.9 | 69 | 5.3 |

**Table 2.** Scan acquisition and image reconstruction parameters.

|  |  |
| --- | --- |
| Scanner model | **Spectral CT 7500** |
| Tube voltage | 100, 120 kVp |
| Rotation time | 0.272 s |
| Collimation | 16x0.625, 128x0.625 mm |
| Noise Index / DRI level | 10, 16, 23 |
| Slice thickness | 2.5 mm |
| Iterative Reconstruction | iDose<sup>4</sup> Level 3 |
| Reconstruction filter | B |
| Reconstructed field of view | 450 mm |
| Image matrix size | 512 x 512 |
| Pixel spacing (in x and y) | 0.88 mm |

### Image analysis

An in-house software tool [14, 39–41] was used to automatically place the regions of interest (ROI) on each insert (covering 60% of the insert diameter, i.e. 17 mm) for MonoE 70 keV images of both insert configurations and phantom size combinations obtained with a DRI of 23. These ROI placements were then copied to corresponding locations on other spectral results to measure the mean and standard deviation of voxel values. Mean and standard deviation from three scans were calculated from multiple central slices from each repeated scan (4 slices x 3 scans at 16×0.625 mm collimation, 10 slices x 3 scans at 128×0.625 mm collimation). Measured values from each of the spectral results from config 1 were compared to the calculated expected range of values. These results were illustrated by plotting HU values for each insert versus keV with an envelope that represents the range of expected values. In addition, for each insert, Levene’s test was utilized to compare the variance of MonoE values from each keV against conventional images across the variation of acquisition parameters and phantom size. A p value of 0.05 was considered significant. Iodine density results for iodine-containing inserts in config 1 were visualized using a scatter plot where error bars corresponded to a single standard deviation of the means.

The stability of MonoE 70 keV relative to conventional images was investigated by determining patient size, tube voltage, radiation dose level, and collimation width dependencies using config 1. Size dependence was calculated as the difference between measured values from the small and large phantom at a DRI of 16. Similarly, tube voltage dependence was calculated as the difference between measured values at 100 and 120 kVp at the moderate dose level while collimation dependence was defined as the difference between measured values at 16×0.625 and 128×0.625 mm. Size and tube voltage dependencies were additionally visualized in scatter plots to demonstrate differences in dependence between conventional and MonoE 70 keV images. For brain, iodine 10 mg/mL, and calcium 100 mg/mL inserts, dose dependency was represented as the range between average mean values for three doses for each tube voltage, collimation, and size combinations. The same calculation was performed for average standard deviation (noise) values.

To highlight material separability and characterization with MonoEs, MonoE 50 keV values were plotted against MonoE 150 keV values for liver-related inserts in config 2. Accuracy of material quantification for the different acquisition parameters and phantom sizes was analyzed by first performing a linear two-base material decomposition utilizing MonoE 50 and 150 keV data. This was achieved by linearly fitting HU values from inserts containing adipose and repeated for inserts containing iron to obtain an origin point that would theoretically correspond to a liver insert. A least squares fit was then applied to the data to obtain material unit vector representations for adipose and iron. Adipose content and iron density for each combination of acquisition parameters and phantom size was estimated from material basis projections for each insert. Both average error and root mean square error (RMSE) across the inserts for each set of acquisition parameters were calculated relative to specified manufacturer values of adipose content and iron density to evaluate material quantification accuracy.

## Results

### Spectral accuracy

MonoEs and iodine maps demonstrated high accuracy for insert measurements across all evaluated energies, phantom sizes, and acquisition parameters. Measured values for each insert mostly fell within the expected range of values as calculated from manufacturer-reported material compositions and physical densities. The largest deviation from the expected attenuation values corresponded to lower keV MonoEs and high density materials (iodine, calcium) (Figure 2). Iodine density demonstrated good accuracy for iodine-containing inserts with an error ranging from -0.25 to 0.5 mg/mL (Figure 3). For both MonoEs and iodine density maps, small differences in measured values were observed amoung different phantom sizes and acquisition parameters. The variations for MonoEs were significantly less than those observed with conventional images (p values ranging from 0.048 to 1.04 × 10^−8^), with the exception of the adipose insert for MonoEs 40 and 50 keV and the brain insert for MonoEs at all keVs except 70 keV.

**Figure 2.**
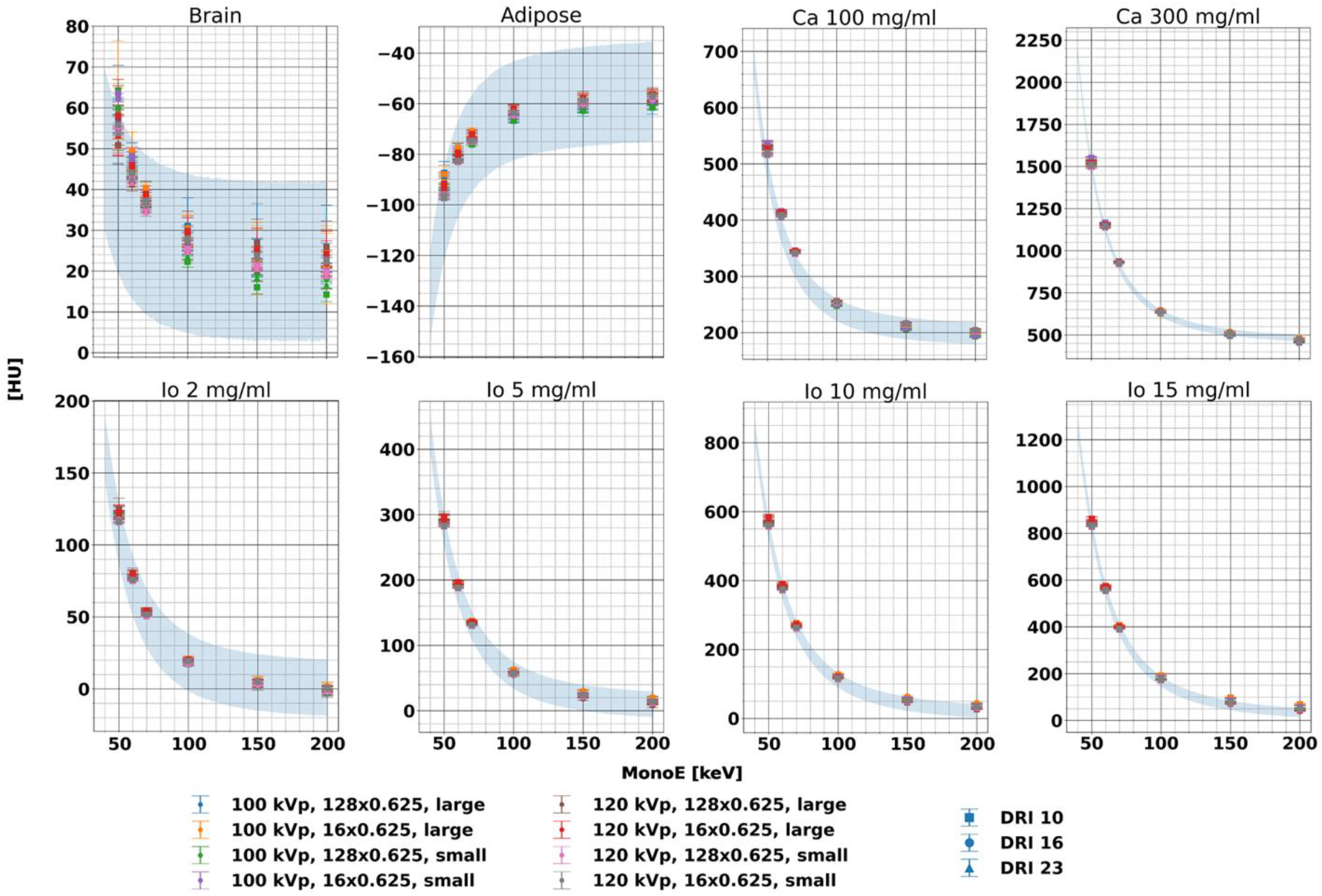
Accuracy of MonoE attenuation for each individual insert and acquisition parameter combination. Measured HU for each insert and keV fell within the range of expected values (shaded blue area) calculated from elemental composition and manufacturer reported physical density, indicating accuracy. Please note different scaling of y-axes.

**Figure 3.**
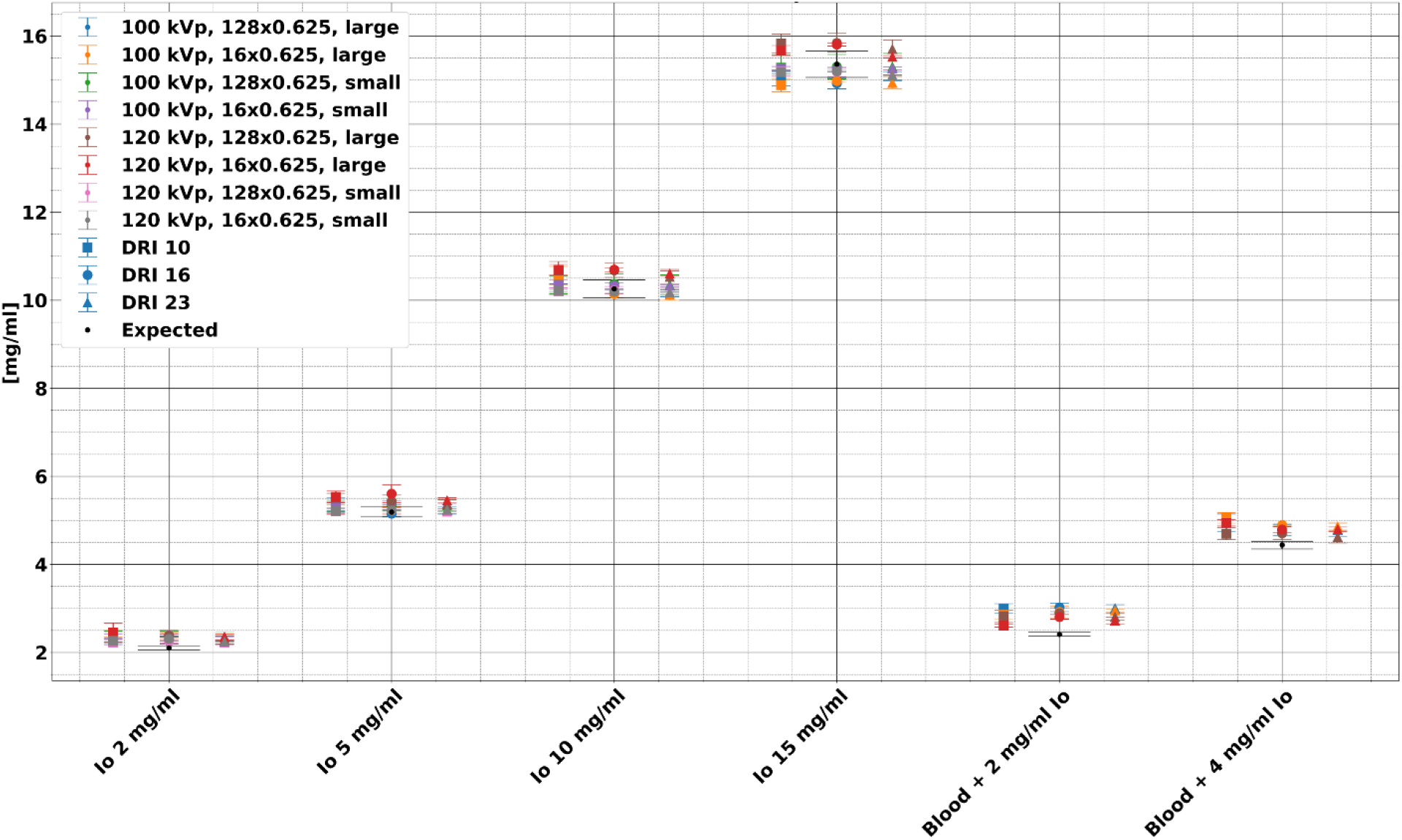
Comparison of measured iodine density for each iodine-containing insert between scans with different phantom sizes, tube voltage, collimation, and dose levels. Measured iodine density was generally accurate and within the expected range of values, with larger deviations for inserts containing blood and iodine. The blood + 2 mg/mL and blood + 4 mg/mL inserts were only available with the large phantom size as the inserts were placed in the extension ring in config 1.

### Size and acquisition parameter dependence

Size and tube voltage dependence of attenuation measurements were dramatically less for MonoE 70 keV in comparison to conventional images, while collimation width dependence maintained a similar low magnitude for both MonoE 70 keV and conventional images. Differences in size resulted in measured differences for inserts ranging from -73.9 to 3.5 HU and -0.6 to 9.9 HU at 120 kVp for conventional and monoE 70 keV images, respectively. At 100 kVp, size dependency ranged from -76.1 to 2.1 HU and -2.7 to 9.5 HU for conventional and MonoE 70 keV images, respectively (Figure 4). In addition, tube voltage dependence of insert attenuation averaged -43.4 and -4.0 HU for conventional and MonoE 70 keV images, respectively, with the small phantom while it averagedv -42.4 and -3.0 HU for conventional and MonoE 70 keV images, respectively, with the large phantom (Figure 5). Both size and tube voltage dependence highlighted the stability of MonoE attenuation in comparison to conventional images that are strongly affected by patient size and acquisition parameters. Collimation dependence (10 vs. 80 mm), on the other hand, did not show a large difference between conventional and MonoE 70 keV images. Measured differences for conventional and MonoE 70 keV images resulting from collimation changes ranged from -9.3 to 0.6 HU and -5.5 to 0.4 HU, respectively. Dose dependence expressed in terms of average mean CT number (Table 3) and average standard deviation (noise) (Table 4) also did not demonstrate a large difference between conventional and MonoE 70 keV images with ranges for noise of 14.8 and 11.9 HU for conventional and MonoE 70 keV images, respectively, with the large phantom. With the small phantom, the ranges of noise were 7.5 and 6.5 HU for conventional and MonoE 70 keV images, respectively.

**Table 3:** Average mean CT number for select inserts for different acquisition parameters.

| Tube Voltage (kVp) | Size | Collimation (mm) | DRI | Average mean CT number [HU] |  |  |  |  |  |
| --- | --- | --- | --- | --- | --- | --- | --- | --- | --- |
|  |  |  |  | Conventional |  |  | MonoE 70 keV |  |  |
|  |  |  |  | Brain | Io 10 mg/mL | Ca 100 mg/mL | Brain | Io 10 mg/mL | Ca 100 mg/mL |
| 100 | large | 128x0.625 | 10 | 34.0 | 291.6 | 360.7 | 40.1 | 276.2 | 344.4 |
|  |  |  | 16 | 36.4 | 293.2 | 358.1 | 38.5 | 272.5 | 342.8 |
|  |  |  | 23 | 36.0 | 291.6 | 356.3 | 36.7 | 271.0 | 342.9 |
|  |  | 16x0.625 | 10 | 37.2 | 295.0 | 363.6 | 40.3 | 276.8 | 345.6 |
|  |  |  | 16 | 39.4 | 297.3 | 362.6 | 40.0 | 274.0 | 345.5 |
|  |  |  | 23 | 37.7 | 296.0 | 360.5 | 38.7 | 273.2 | 346.3 |
|  | small | 128x0.625 | 10 | 37.6 | 334.8 | 385.4 | 36.3 | 266.7 | 341.7 |
|  |  |  | 16 | 36.5 | 334.8 | 385.8 | 36.4 | 267.4 | 342.2 |
|  |  |  | 23 | 36.7 | 335.0 | 385.6 | 35.4 | 267.0 | 341.2 |
|  |  | 16x0.625 | 10 | 39.3 | 338.7 | 388.2 | 38.9 | 269.7 | 343.9 |
|  |  |  | 16 | 38.7 | 338.7 | 388.2 | 38.4 | 269.8 | 343.8 |
|  |  |  | 23 | 38.2 | 338.7 | 388.2 | 37.1 | 269.8 | 343.2 |
| 120 | large | 128x0.625 | 10 | 32.4 | 232.4 | 325.5 | 36.8 | 270.5 | 343.6 |
|  |  |  | 16 | 31.1 | 233.6 | 325.1 | 34.6 | 268.5 | 342.3 |
|  |  |  | 23 | 31.7 | 232.1 | 322.7 | 35.2 | 265.9 | 340.6 |
|  |  | 16x0.625 | 10 | 33.9 | 236.3 | 328.2 | 38.9 | 273.3 | 345.4 |
|  |  |  | 16 | 34.3 | 236.7 | 327.9 | 37.8 | 269.9 | 344.7 |
|  |  |  | 23 | 32.8 | 235.0 | 325.8 | 36.9 | 267.1 | 343.5 |
|  | small | 128x0.625 | 10 | 33.2 | 274.7 | 349.6 | 35.5 | 261.3 | 340.4 |
|  |  |  | 16 | 32.4 | 274.6 | 349.3 | 35.2 | 261.1 | 340.1 |
|  |  |  | 23 | 32.3 | 274.6 | 349.5 | 34.5 | 260.8 | 339.6 |
|  |  | 16x0.625 | 10 | 34.3 | 277.9 | 351.8 | 37.2 | 263.4 | 342.0 |
|  |  |  | 16 | 33.8 | 277.6 | 351.5 | 36.9 | 263.5 | 341.7 |
|  |  |  | 23 | 33.6 | 277.8 | 351.7 | 36.1 | 263.3 | 341.2 |

**Table 4:** Average standard deviation of CT number for select inserts for different acquisition parameters.

| Tube Voltage (kVp) | Size | Collimation (mm) | DRI | Average standard deviation of CT number [HU] |  |  |  |  |  |
| --- | --- | --- | --- | --- | --- | --- | --- | --- | --- |
|  |  |  |  | Conventional |  |  | MonoE 70 keV |  |  |
|  |  |  |  | Brain | Io 10 mg/mL | Ca 100 mg/mL | Brain | Io 10 mg/mL | Ca 100 mg/mL |
| 100 | large | 128x0.625 | 10 | 38.7 | 40.0 | 37.1 | 31.7 | 32.5 | 30.2 |
|  |  |  | 16 | 33.6 | 32.9 | 29.9 | 27.9 | 26.9 | 24.9 |
|  |  |  | 23 | 25.9 | 25.1 | 22.6 | 21.8 | 20.6 | 18.8 |
|  |  | 16x0.625 | 10 | 40.3 | 37.6 | 35.4 | 32.8 | 30.6 | 29.0 |
|  |  |  | 16 | 32.8 | 31.4 | 28.7 | 27.0 | 25.5 | 23.6 |
|  |  |  | 23 | 26.1 | 24.7 | 22.8 | 21.9 | 20.5 | 18.9 |
|  | small | 128x0.625 | 10 | 17.2 | 14.5 | 14.9 | 14.6 | 11.9 | 12.3 |
|  |  |  | 16 | 13.3 | 11.8 | 11.9 | 11.1 | 9.4 | 9.7 |
|  |  |  | 23 | 8.0 | 7.3 | 7.7 | 6.7 | 5.5 | 5.8 |
|  |  | 16x0.625 | 10 | 16.9 | 14.1 | 14.3 | 14.3 | 11.6 | 11.8 |
|  |  |  | 16 | 13.2 | 11.3 | 11.4 | 11.1 | 9.2 | 9.4 |
|  |  |  | 23 | 7.9 | 6.9 | 7.0 | 6.5 | 5.4 | 5.6 |
| 120 | large | 128x0.625 | 10 | 34.0 | 32.5 | 31.0 | 28.4 | 27.2 | 26.1 |
|  |  |  | 16 | 27.8 | 26.8 | 26.1 | 23.3 | 22.5 | 22.0 |
|  |  |  | 23 | 16.3 | 16.2 | 16.0 | 13.9 | 13.9 | 13.5 |
|  |  | 16x0.625 | 10 | 34.1 | 32.2 | 30.4 | 28.3 | 26.7 | 25.2 |
|  |  |  | 16 | 28.9 | 26.8 | 24.9 | 24.2 | 22.4 | 20.8 |
|  |  |  | 23 | 17.1 | 17.0 | 15.8 | 14.5 | 14.3 | 13.4 |
|  | small | 128x0.625 | 10 | 15.5 | 13.9 | 13.4 | 13.2 | 11.4 | 11.0 |
|  |  |  | 16 | 12.2 | 11.2 | 11.5 | 10.2 | 8.8 | 9.5 |
|  |  |  | 23 | 7.4 | 7.0 | 6.9 | 6.1 | 5.1 | 5.4 |
|  |  | 16x0.625 | 10 | 15.3 | 13.0 | 13.3 | 12.9 | 10.7 | 11.0 |
|  |  |  | 16 | 12.3 | 10.7 | 10.6 | 10.3 | 8.7 | 8.7 |
|  |  |  | 23 | 7.4 | 6.5 | 6.5 | 6.1 | 5.0 | 5.2 |

**Figure 4.**
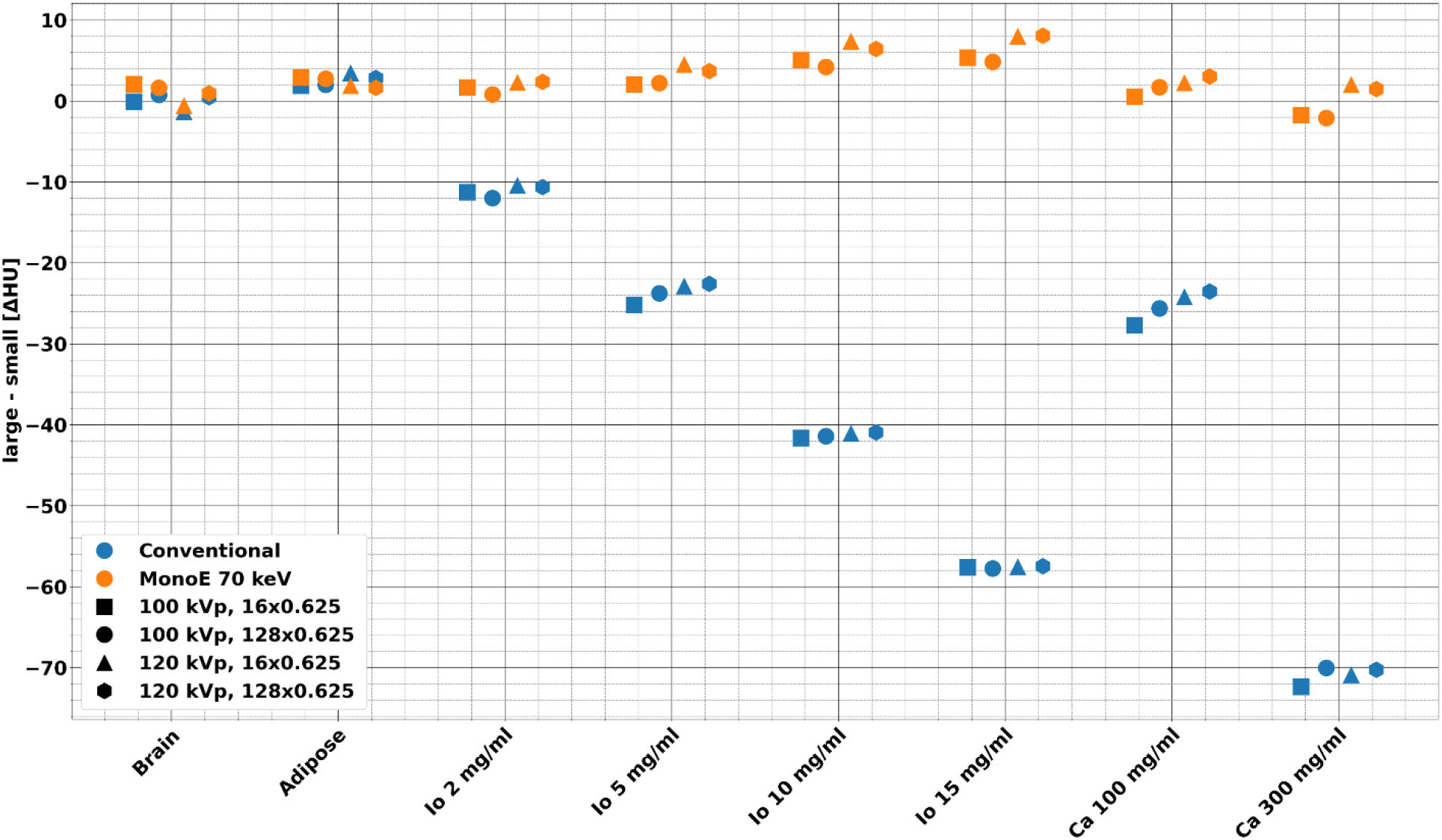
Size dependence of conventional and MonoE 70 keV images for each individual insert at DRI 16. The differences between the large and small phantoms were less evident in MonoE 70 keV compared to conventional images. This trend applied to all inserts and was particularly clear with higher density materials.

**Figure 5.**
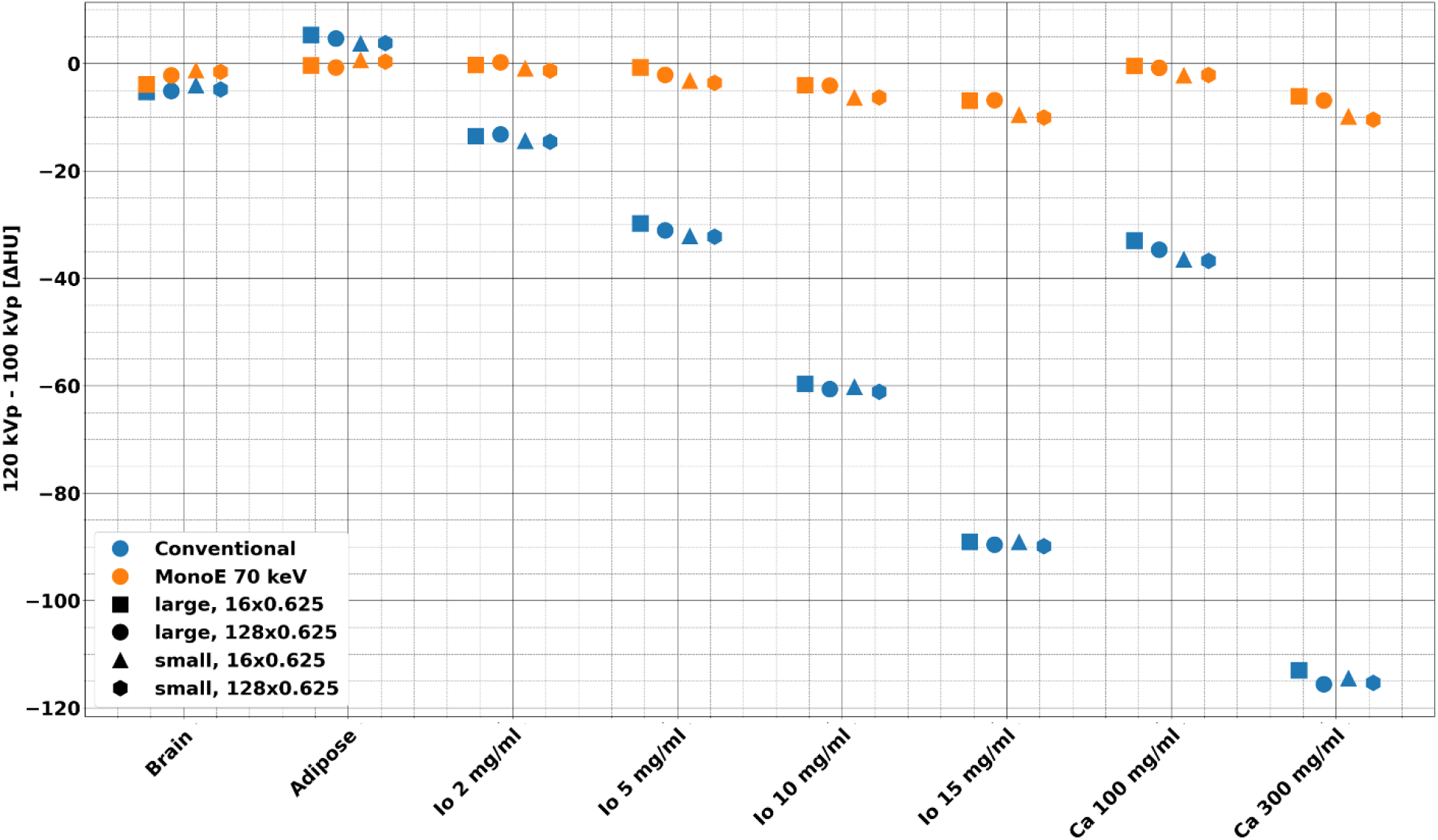
Differences in CT numbers for conventional images and MonoE 70 keV between 100 and 120 kVp at DRI 16. MonoE 70 keV demonstrated low tube voltage dependence in comparison to conventional images with differences in the large phantom ranging from -11 to 1 HU and -116 to 7 HU for MonoE 70 keV and conventional images, respectively. For the small phantom, tube voltage dependence ranged from -11 to 1 HU and -115 to 4 HU for MonoE 70 keV and conventional images, respectively.

### Material characterization

Figure 6 highlights material characterization and quantification capabilities with MonoE 50 and 150 keV for inserts mimicking liver with different adipose content and iron concentration. Despite small variations resulting from different phantom size and acquisition parameter combinations, values from the same insert aggregated around a single pair of MonoE 50 and 150 keV values but remained separated from values from inserts of different composition, showing excellent material characterization and separability. Moreover, both adipose-containing and iron-containing inserts demonstrated a linear relationship that corresponds to the material content. Based on specified manufacturer values, mean RMSE (Table 5) for adipose content and iron density across inserts averaged 1.78 ± 0.37% (range: 0.47% - 3.36%) and 0.87 ± 0.37 mg/mL (range: 0.25 mg/mL – 1.76 mg/mL) for acquisition parameter combinations, respectively, demonstrating material quantification accuracy. Additionally, average relative error across inserts ranged from -4.4 to 5.0% for adipose content and from -2.2 to 2.6 mg/mL for iron concentration (Table 5) for different acquisition parameter and phantom size combinations.

**Table 5.** Average relative error and root mean square error (RMSE) of adipose content and iron concentration quantification for different acquisition parameters.

| Tube Voltage (kVp) | Size | Collimation (mm) | DRI | Average relative error |  | RMSE |  |
| --- | --- | --- | --- | --- | --- | --- | --- |
|  |  |  |  | Adipose [%] | Iron [mg/mL] | Adipose [%] | Iron [mg/mL] |
| 100 | large | 128x0.625 | 10 | 1.46 | 1.04 | 1.85 | 1.16 |
|  |  |  | 16 | 0.38 | 0.45 | 1.19 | 0.52 |
|  |  |  | 23 | -0.80 | -0.11 | 1.32 | 0.52 |
|  |  | 16x0.625 | 10 | 5.04 | 2.62 | 3.35 | 1.76 |
|  |  |  | 16 | 3.79 | 1.70 | 2.76 | 1.14 |
|  |  |  | 23 | 4.89 | 2.14 | 3.36 | 1.48 |
|  | small | 128x0.625 | 10 | 1.58 | 0.89 | 1.15 | 0.63 |
|  |  |  | 16 | 1.63 | 0.91 | 1.14 | 0.62 |
|  |  |  | 23 | 0.28 | 0.17 | 0.47 | 0.25 |
|  |  | 16x0.625 | 10 | 4.05 | 1.63 | 2.61 | 1.04 |
|  |  |  | 16 | 3.18 | 1.27 | 2.02 | 0.81 |
|  |  |  | 23 | 1.87 | 0.59 | 1.30 | 0.46 |
| 120 | large | 128x0.625 | 10 | -1.76 | -0.62 | 1.23 | 0.57 |
|  |  |  | 16 | -2.88 | -1.25 | 1.91 | 0.88 |
|  |  |  | 23 | -4.39 | -2.17 | 2.83 | 1.40 |
|  |  | 16x0.625 | 10 | 0.78 | 0.64 | 1.28 | 0.89 |
|  |  |  | 16 | 0.01 | 0.17 | 0.84 | 0.45 |
|  |  |  | 23 | -2.14 | -1.13 | 1.68 | 0.86 |
|  | small | 128x0.625 | 10 | -3.36 | -1.57 | 2.16 | 1.01 |
|  |  |  | 16 | -2.96 | -1.40 | 1.94 | 0.90 |
|  |  |  | 23 | -4.31 | -2.11 | 2.74 | 1.33 |
|  |  | 16x0.625 | 10 | -1.47 | -0.96 | 1.19 | 0.68 |
|  |  |  | 16 | -1.07 | -0.82 | 0.91 | 0.56 |
|  |  |  | 23 | -2.34 | -1.49 | 1.54 | 0.94 |

**Figure 6.**
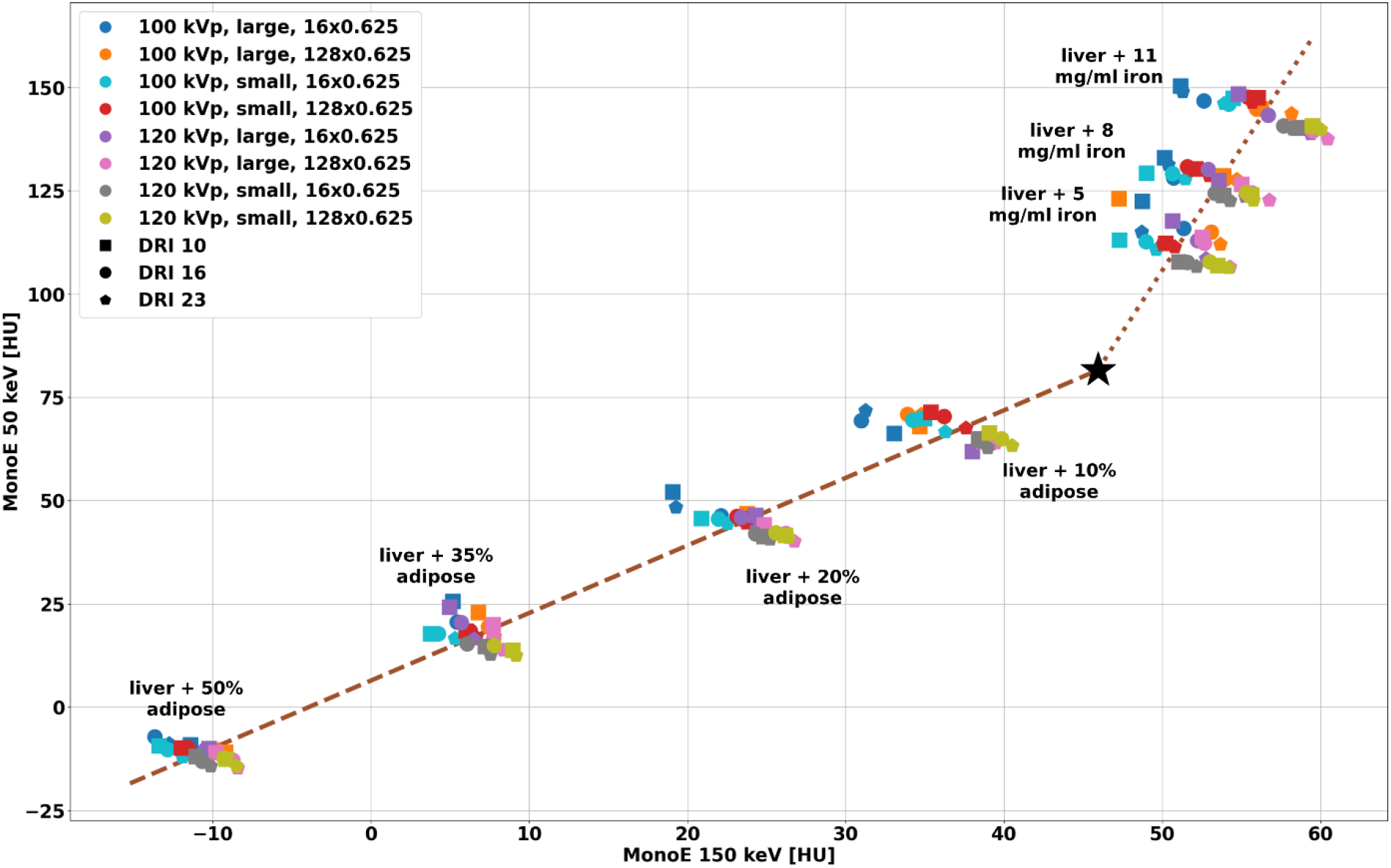
Separation of materials with MonoE 50 and 150 keV for config 2. CT numbers measured for MonoE 50 and 150 keV for the same insert exhibited small variations with different tube voltages, phantom size, collimation width, and dose levels. Values from the same insert aggregated and were distinct from other inserts. The origin (black star) and vectors for adipose content (brown dashed line) and iron density (brown dotted line) from the linear two-base material decomposition are also shown. The results indicated the ability to quantify adipose content and iron concentration.

## Discussion

In this study, our objective was to characterize the spectral imaging performance of a second-generation dual-layer spectral CT scanner. We demonstrated several benefits of spectral CT, including **(i)** a decoupling of image quality from acquisition parameters (100 versus 120 kVp, 10 versus 80 mm z-coverage) and patient habitus, **(ii)** accurate quantitative imaging, especially in the case of iodine, with minimal impact from collimation, radiation dose levels, and patient habitus, and **(iii)** excellent material characterization capabilities, such as for iron.

Compared to first-generation dual-layer spectral CT, iodine quantification with the second-generation dual-layer spectral CT was improved. Utilizing the same multi-energy CT phantom, prior studies with a first-generation dual-layer spectral CT demonstrated an iodine bias [16] of 1.03 mg/mL and absolute error [42] of 0.5 mg/mL at 120 kVp with the large phantom. Our study, however, demonstrated an improved iodine bias of 0.86 mg/mL and absolute error of 0.31 mg/mL at 120 kVp. Similarly, at 100 kVp, the iodine bias was small, at -0.12 mg/mL, as was the absolute error at 0.23 mg/mL, highlighting improved iodine quantification. The previous system did not provide a 100 kVp scanning option. As a result of these improvements in iodine quantification with the second generation of systems, spectral CT may offer advantages over conventional oncological therapy assessment by using a biomarker that is based on iodine density. By utilizing iodine concentration measurements, one can obtain a functional image of perfusion patterns, which can be used to link biomarkers to changes in lesion shape, topology, texture, and vasculature [35–38] more accurately than the current oncological assessment criteria [36, 43, 45]. Furthermore, by extending the maximum z-coverage from 4 cm to 8 cm, iodine quantification in the myocardium may be used as a robust, semi-quantitative proxy for myocardial perfusion, with iodine concentration serving as an indicator of blood distribution at a given time [17]. To this end, we have presented results in this study which demonstrate that collimation width has minimal effect on iodine quantification. Additionally, spectral CT can be used to characterize materials outside the iodine domain, including iron [44, 46, 47]. As spectral CT technology improves, as described in this study, it is becoming increasingly possible to characterize and detect iron content. This development may have clinical implications beyond liver imaging to include cardiac diagnostics as well [48].

There are limitations to the present study. First, this study characterized the system by using technical phantoms and not clinical patient data. There will be a need to perform quantitative and qualitative disease-specific studies in the future to determine performance across the diagnostic range. However, using phantoms, we have knowledge of ground-truth material compositions and densities, which is not available in clinical trials. Additionally, despite the fact that the investigated system advances cardiac imaging with an extended z-coverage, we did not investigated the effect of heart rate on spectral performance. In a follow-up study, we intend to investigate the relationship between temporal resolution and spectral image quality. Unlike previous studies of dual-layer CT scanners [49], we have not included experiments at 140 kVp, but rather have focused on 100 kVp and 120 kVp. In this regard, 100 kVp is clearly more suitable for smaller patients, especially pediatric patients [41]. Due to the smaller habitus of children, the use of lower tube voltages and dedicated CT protocols can reduce radiation exposure, and this study demonstrates the ability to do so without compromising spectral performance. Finally, our study does not include direct comparisons with other spectral CT platforms. We and other investigators have described the performance characteristics of other platforms, including photon-counting, in previous publications [13–17].

## Conclusion

To conclude, we report the results of an experimental evaluation with respect to spectral accuracy, patient size and acquisition parameter dependence, and material characterization of a second-generation dual-layer spectral CT. These results demonstrate improvements over first-generation dual-layer spectral CTs, which may improve the clinical value of spectral imaging and lead to more routine clinical adoption. As detector-based spectral CT continues to demonstrate improved performance over conventional CT, its additional diagnostic benefits will become an expected part of routine clinical evaluation.

## Data Availability

All data produced in the present study are available upon reasonable request to the authors

## Abbreviations

CT: computed tomography
HU: Hounsfield units
DECT: dual-energy CT
MonoE: virtual monoenergetic images
DRI: dose right index
CTDI_vol_: CT dose index
ROI: region of interest
RMSE: root mean square error

## Acknowledgments

The authors acknowledge support through the National Institutes of Health (R01EB030494). In addition, the authors would like to thank Safaa Abdallah for her help with acquiring data.

## Conflict of Interest

Harold I. Litt and Peter B. Noël received a hardware grant from Philips Healthcare. Peter B. Noël receives research grant funding from Philips Healthcare. Tim Leiner, Peter B. Noël, and Wolfram Stiller are members of the CT Advisory Board of Philips Medical Systems. Hans-Ulrich Kauczor receives research grant funding and speakers bureau fees from Philips Healthcare. Sandra Halliburton and Amy Perkins are employees of Philips Healthcare. The other authors have no relevant conflicts of interest to disclose.

## Notes

### Funding Statement

This study was funded by the National Institutes of Health (R01EB030494).

